# STIMULATE-ICP: A pragmatic, multi-centre, cluster randomised trial of an integrated care pathway with a nested, Phase III, open label, adaptive platform randomised drug trial in individuals with Long COVID: a structured protocol

**DOI:** 10.1101/2022.07.21.22277893

**Authors:** Denise Forshaw, Emma C Wall, Gordon Prescott, Hakim-Moulay Dehbi, Angela Green, Emily Attree, Lyth Hismeh, William D Strain, Michael G Crooks, Caroline Watkins, Chris Robson, Rajarshi Banerjee, Paula Lorgelly, Melissa Heightman, Amitava Banerjee, the STIMULATE-ICP trial team

## Abstract

**Introduction:** Long COVID (LC), the persistent symptoms ≥12 weeks following acute COVID-19, presents major threats to individual and public health across countries, affecting over 1.5 million people in the UK alone. Evidence-based interventions are urgently required and an integrated care pathway (ICP) approach in pragmatic trials, which include investigations, treatments and rehabilitation for LC, could provide scalable and generalisable solutions at pace.

**Methods and analysis:** This is a pragmatic, multi-centre, cluster-randomised clinical trial of two components of an ICP (Coverscan^™^, a multi-organ MRI, and Living with COVID Recovery^™^, a digitally enabled rehabilitation platform) with a nested, Phase III, open label, platform randomised drug trial in individuals with LC. Cluster randomisation is at level of primary care networks so that ICP interventions are delivered as “standard of care” in that area. The drug trial randomisation is at individual level and initial arms are rivaroxaban, colchicine, famotidine/loratadine, compared with no drugs, with potential to add in further drug arms. The trial is being carried out in 6-10 NHS LC clinics in the UK and is evaluating the effectiveness of a pathway of care for adults with LC in reducing fatigue and other physical, psychological and functional outcomes (e.g. EQ-5D-5L, GAD-7, PHQ-9, WSAS, PDQ-5, CFQ, SF-12, MRC Dyspnoea score) at 3 months. The trial also includes an economic evaluation which will be described separately.

**Ethics and dissemination:** The protocol was reviewed by South Central - Berkshire Research Ethics Committee (reference: 21/SC/0416). All participating sites obtained local approvals prior to recruitment. Coverscan^™^ has UKCA certification (752965). The first participant was recruited in July 2022 and interim/final results will be disseminated in 2023, in a plan co-developed with public and patient representatives. The results will be presented at national and international conferences, published in peer reviewed medical journals, and shared via media (mainstream and social) and patient support organisations.

**Trial registration number:** ISRCTN10665760

## Introduction

The syndrome of Long COVID (LC), defined by persistent post-COVID-19 symptoms ≥12 weeks(1), has affected over 2 million people in the UK alone(2), necessitating health system responses whilst research is still defining the disease. While many symptoms are reported by individuals with LC, fatigue is the most frequent symptom and is most commonly reported as the symptom that most limits quality of life (3)(4). Among other symptoms reported are chest pain, cognitive slowing, reported commonly as ‘brain fog’, dyspnoea, headache, dizziness, palpitations, and sleep disturbances (5). Unlike acute COVID-19(6), predictors of poor outcomes and effective treatments are not yet established for LC(7,8), which spans multiple healthcare challenges, particularly how to deliver sustainable, high-quality care for multimorbidity and long-term conditions (LTC)(9). Integrated care pathways (ICP) are structured, multidisciplinary plans of essential steps in care of specific conditions(10). An ICP approach offers coordination across specialties, investigations, treatment, and rehabilitation, as well as opportunities for real-time, iterative improvements in service design and delivery(11–13).

There is variation in access to referral and care, and poor patient experience for LC(14). With stretched resources during the pandemic, rationalisation of investigation and rehabilitation across diseases is crucial. In LC, non-hospitalised individuals were more symptomatic, more likely to have psychological impact, and less likely to be fit for work than post-hospitalised individuals. In LC, where up to 70% of individuals show evidence of mild impairment in ≥1 organs(15), an early supported investigation strategy may direct management, as well as better defining the syndrome, with relevance to other LTCs. There is no evidence-based treatment or rehabilitation, with few clinical trials to-date, particularly in non-hospitalised individuals, or in the context of overall care pathways. Despite roll-out of 90 centrally funded LC clinics in England, ICPs are neither coordinated nor evaluated. Trials of ICPs could inform diagnosis, care, public health, policy planning, resource allocation and budgeting.

Antihistamines (e.g., loratadine and famotidine), colchicine and rivaroxaban are examples of medications being widely used among patients and health professionals with rationale for use (**Appendix 1**), but without proven efficacy, necessitating trials. Platform trials have been used at scale to evaluate therapies in acute COVID-19(16), but not yet in LC. Since the underlying causes of LC are unknown and subtypes are poorly defined, trials should test drugs across all individuals with LC, irrespective of their individual symptoms or investigation findings, to allow multiple secondary sub-group analyses. The optimal outcomes for LC are yet to be fully defined. Therefore, trials should evaluate a wide range of potential physical, psychological and functional outcomes. We report the protocol for the STIMULATE-ICP (Symptoms, Trajectory, Inequalities and Management: Understanding Long COVID to Address and Transform Existing Integrated Care Pathways) trial (**Figure 1**).

**Figure 1.**
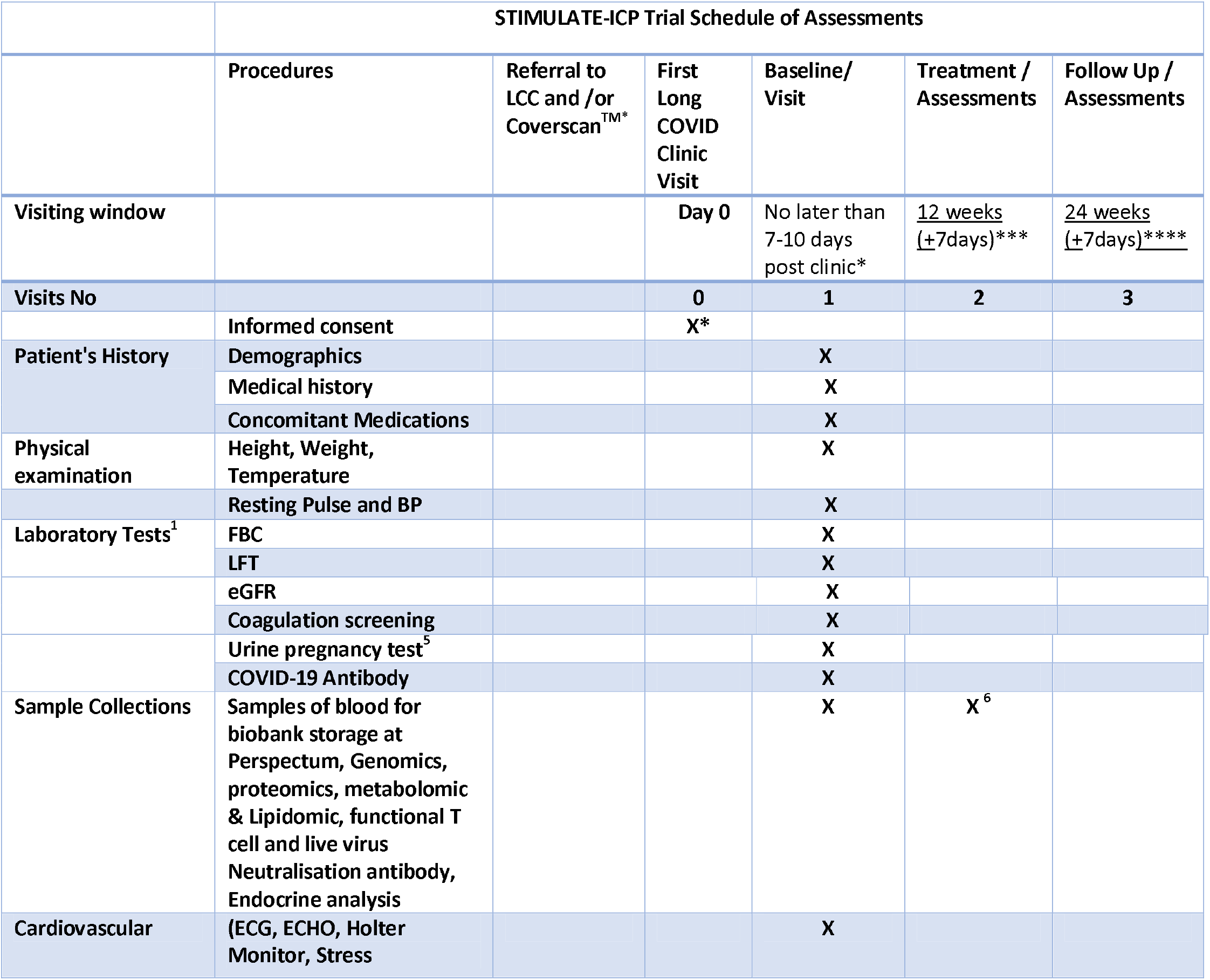
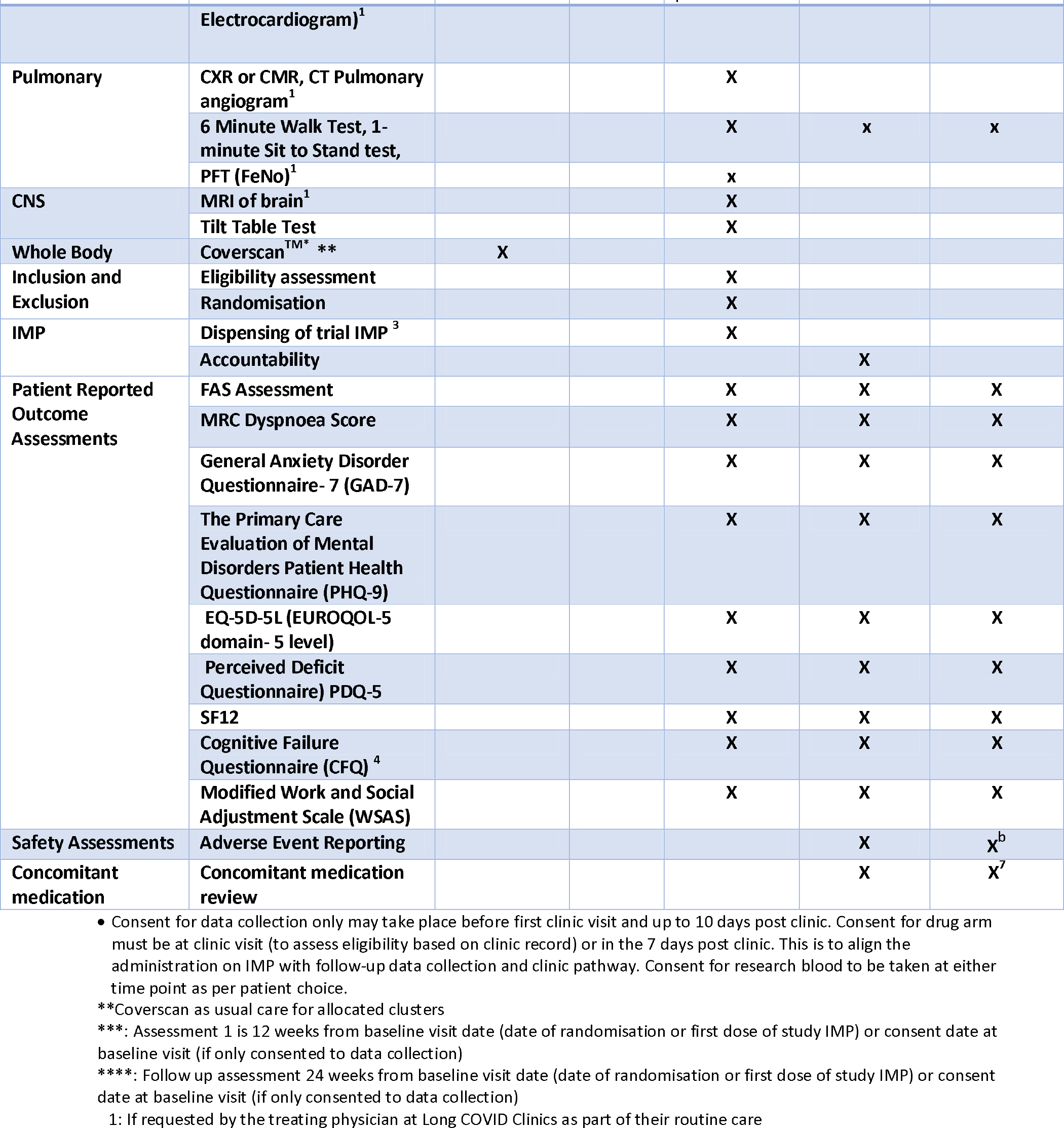
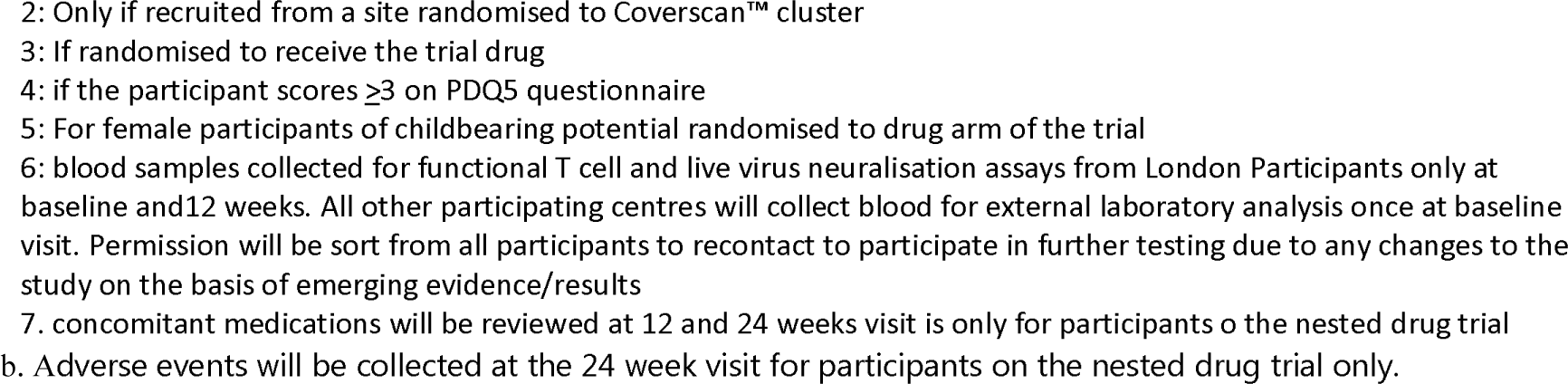
Schedule of Assessments.

### Objectives

For individuals with LC, to evaluate the:

1. effect of “integrated care” versus “usual care”.
2. clinical efficacy of
  a. individual components of an ICP
  b. potential drug therapies within a nested drug trial.
3. pathophysiology, trajectory and outcomes, including healthcare utilisation (**Table 1 and Table 2**)

**Table 1:** Research question in STIMULATE-ICP.

|  |
| --- |
| **Over-arching research question:**<br>What is the effectiveness of an integrated care pathway for individuals with Long Covid, compared with usual care? |
| **Population:**<br>Individuals $\geq 18$ years with $\geq 4$ weeks post-COVID symptoms (in line with UK clinical guidance and policy), based on clinician diagnosis, newly referred to participating Long COVID clinics. |
| **Intervention:**<br>*Investigation:* Multi-organ MRI prior to Long Covid clinic assessment (Coverscan™)<br>*Treatment:* Multiple drug arms, starting with famotidine/loratadine, colchicine and rivaroxaban<br>*Rehabilitation:* Digitally enabled rehabilitation (Living with COVID Recovery™) |
| **Comparator:**<br>*Investigation:* Usual care<br>*Treatment:* No drug<br>*Rehabilitation:* Usual care |
| **Outcomes:**<br>*Primary-* Fatigue on the fatigue assessment scale at 12 weeks.<br>*Secondary-* Multiple assessments of health-related quality of life, mental health (including depression), work and social adjustment, physical function, organ impairment and healthcare utilisation at 12 and 24 weeks. |

**Table 2:**
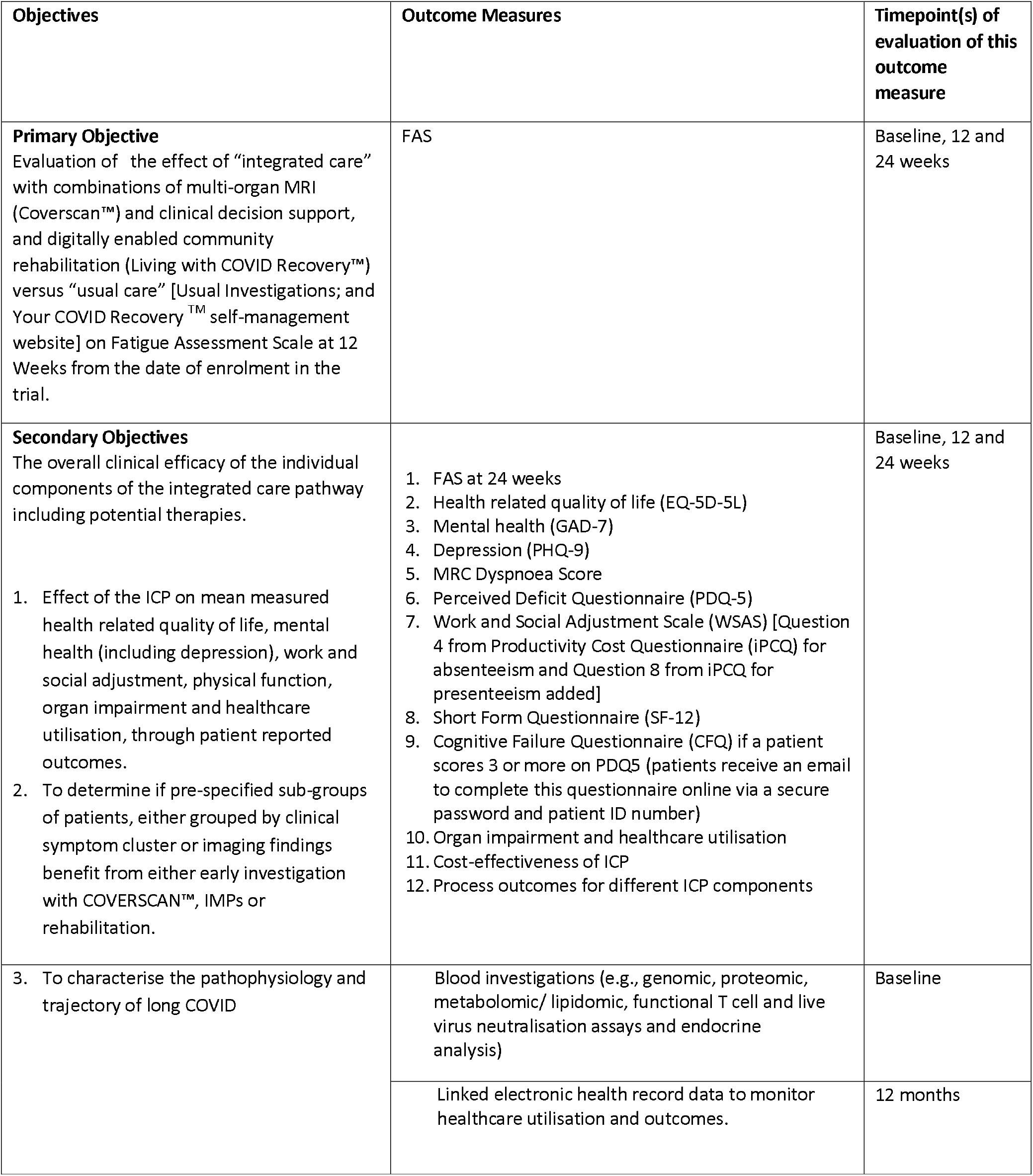
Summary of the primary and secondary objectives and outcome measures.

## Methods and analysis

### Design

STIMULATE-ICP is a pragmatic, cluster-randomised trial of two components of an ICP with a nested, Phase III, open label platform randomised drug trial in individuals with LC. It is responsive to local resources and needs, *reactive* to changing pandemic and policy contexts and *iterative*, depending on ongoing results, as part of a National Institute for Health Research (NIHR: COV-LT2-0043(17)) -funded programme (**Figure 2**), with epidemiologic and mixed methods studies, including care inequalities and transferability to other LTCs(18); IRAS 303958(19)) and this complex intervention trial (IRAS 1004698).

**Figure 2.**
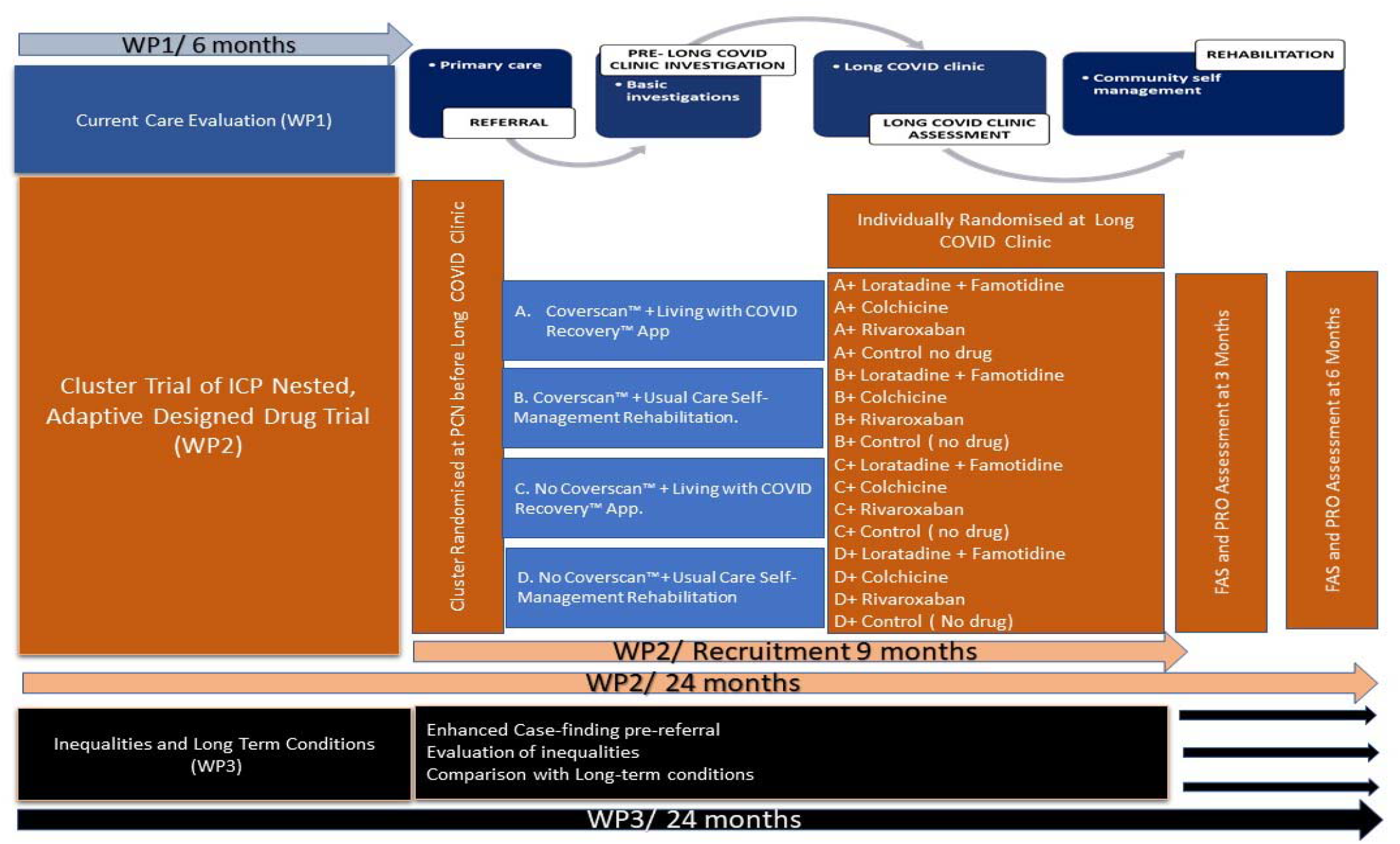
The STIMULATE-ICP programme showing the interconnections between the Trial and other work packages

**Figure 3:**
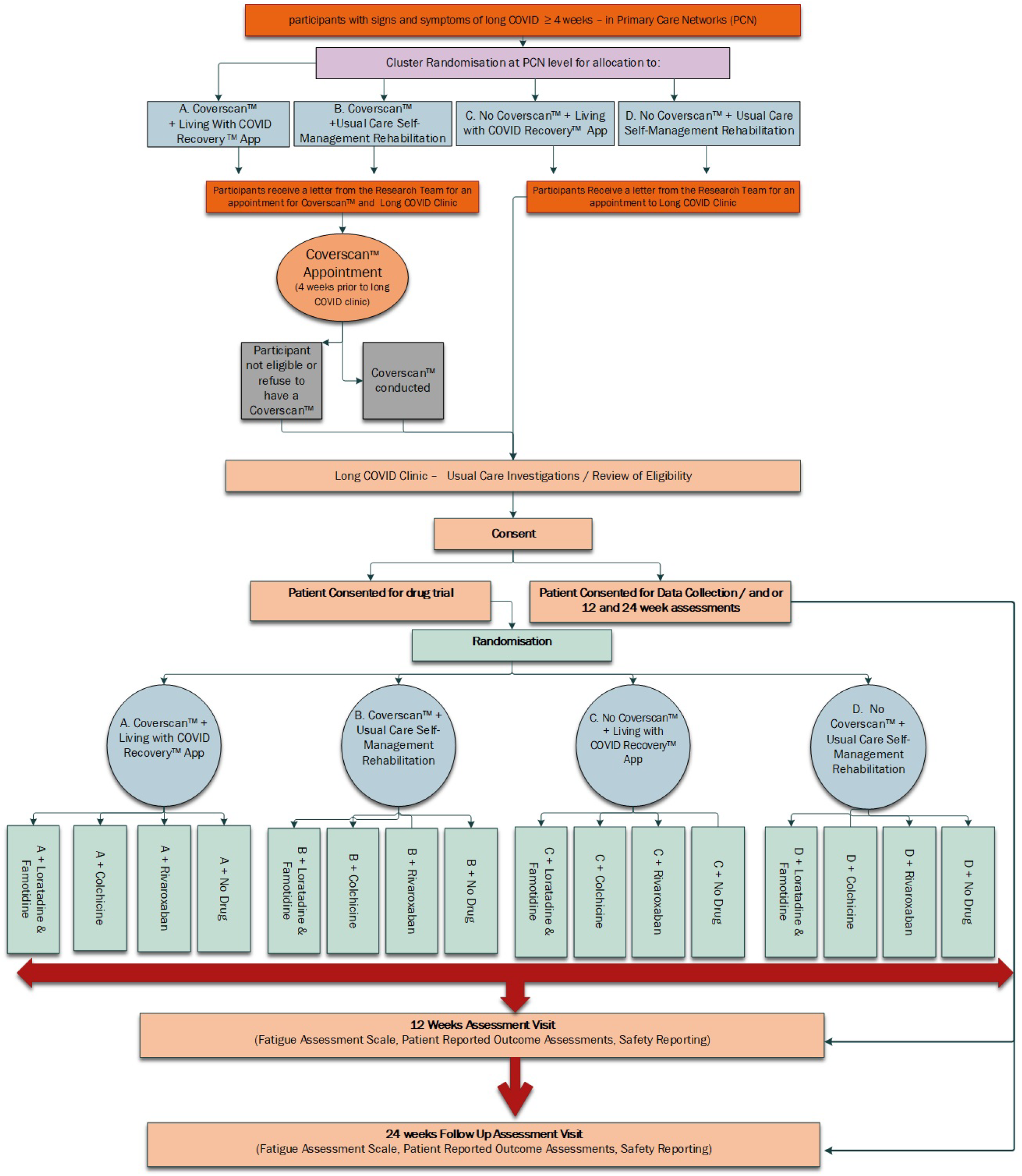
STIMULATE-ICP Study Flow.

### Setting

This is a multi-centre trial involving a combination of Primary Care Networks and LC Clinics in 6-10 different areas of England (depending on throughput and size), purposefully selected to encompassing geography, demographic diversity and design of clinical service delivery. Management is by Lancashire Clinical Trials Unit (LCTU) and sponsored by University College London (UCL).

### Intervention

In this trial, “integrated care” comprises two ICP interventions, cluster-randomised at t he Primary Care Network (PCN) level: Coverscan^™^ (community based, multi-organ MRI with clinical decision support)(15) and Living with COVID Recovery^™^ (enhanced community based, digitally enabled, rehabilitation)(20). “Usual care” comprises usual investigations (e.g. routine blood tests, ECG, chest radiograph and exercise tolerance test); and self-managed rehabilitation with online resources (Your COVID Recovery^™^: https://www.yourcovidrecovery.nhs.uk) in the LC clinic pathway, which is standard of care within individual services.

In a nested, open label, adaptive platform trial, a 12-week course of one of three drug arms will be initially evaluated (individual level randomisation): famotidine+loratadine, colchicine and rivaroxaban, compared with no drugs. Other drugs may be added as additional or replacement arms for new patients at pre-determined time points during the trial (4 and 8 months), dependent on interim trial data analysis with consideration of updated clinical and policy recommendations.

### Sites

This is a pragmatic randomised clinical trial to compare two components of ICPs to usual care, ensuring that research activities do not impact/change workload for already over-stretched NHS resources, whilst ensuring equity of access to care. LC clinic sites will be purposefully selected, depending on throughput and size to encompass diversity of geography, clinic design and demography.

### Randomisation

The treatment pathways (the ICP interventions) are cluster randomised by PCN. Given current COVID-related health system pressures, individual level randomisation for Coverscan^™^ and Living with COVID Recovery^™^ at General Practitioner level is not feasible and could potentially increase waiting times in other areas of care, which could affect recruitment during trial timelines. Unlike individual randomisation, cluster randomising at PCN level allows 50-50 chance of allocation to both Coverscan^™^ and Living with COVID Recovery^™^, maintaining access across the total number of individuals referred to the participating clinic, keeping equipoise at clinic level. Randomisation will use demographic and socioeconomic data available for each PCN, ensuring equity of access through deprivation indexes of GP postcodes. Clustering at PCN level allows Coverscan^™^ and/or Living with COVID Recovery^™^ to be provided as uplift to usual care in allocated PCNs, becoming integrated care within that PCN without the need for individual consent. Access to and collection of Coverscan^™^ and Living with COVID Recovery^™^ data for research will be by individual consent on study entry (by the research staff). Cluster randomisation (2×2 factorial design) provides four groups:

a. Coverscan^™^ + Living with COVID Recovery^™^ App.
b. Coverscan^™^ + Usual Care Self-Management Rehabilitation.
c. No Coverscan^™^ + Living with COVID Recovery^™^ App.
d. No Coverscan^™^+ Usual Care Self-Management Rehabilitation.

At the LC clinic, individuals eligible for the drug study will be randomised at the individual level to one of the following 4 arms:

1. Famotidine + Loratadine
2. Colchicine
3. Rivaroxaban
4. No Drug

Other drugs may be added as additional or replacement arms for new patients at predetermined time points during the trial (6 months and 12 months), dependent on interim analysis of trial data with consideration given to updated recommendations from an independent advisory board.

Individuals who are ineligible for the drug study will invited to participate for data collection. Including these individuals will enable data collection to represent the overall LC population.

### Randomisation Methods

Randomisation to the cluster trial will be at the PCN level. Block randomisation for the PCNs will be used with blocks of size 4. There will be two factors in the randomisation, and within each clinic: PCN size (categorised into 2 levels) and PCN deprivation (categorised into 2 levels). Within each clinic, the strata of PCNs will be arranged as larger and more deprived; larger and less deprived; smaller and less deprived; and smaller and more deprived, before randomised allocation into the 4 cluster allocations. The randomisation ratio is 1:1:1:1 between the 4 possible treatment pathways.

Randomisation to the drug trial at the clinic will be at the individual participant level using an online system provided by Sealed Envelope^™^. Block randomisation will be used with varying block sizes. There will be two factors in the randomisation within each clinic: gender (male vs female) and treatment pathway. The randomisation ratio will be equal between all of the drug treatment arms (including no drug (usual care)) open to recruitment at that point in the cluster trial.

### Timeline

We aim to recruit individuals over a 12-month period. The 12-week treatment duration in the drug trial and the 12-week primary outcome, fatigue, were selected to match the 12-week duration of the commissioned NHS LC clinic pathway. Outcomes will also be assessed at 24 weeks. The overall trial will run over an 18-month period.

### Participants

Participants are ≥18 years with ≥4 weeks post-COVID symptoms (in line with UK clinical guidance and policy), based on clinician diagnosis, newly referred to participating LC clinics. The vast majority of people included in the trial will have had symptoms for >12 weeks based on delays in the current care pathways and our preliminary UCLH data. Inclusion and exclusion criteria are listed below.

*Inclusion Criteria for ALL Participants*

- Capable of giving informed consent.
- Age 18 years and above
- Clinical Parameters: persistent signs and symptoms for a period of 4 weeks or longer in duration post-COVID-19 infection (either by test result or symptomology). Presenting at their first referral first visit to a participating LC clinic pathway.
- Able to read or understand English or have a relative/family member able to read/understand English to facilitate participation (essential for patient reported outcome measures at follow-up time points and virtual contact).
- Not enrolled in any other interventional study where study intervention/activities may affect outcome measures (individuals enrolled in purely observational studies can be included)

Additional Inclusion Criteria for the nested, platform randomised drug trial to be met in addition to **all above criteria** are listed in **Appendix 2**.

### Assessment of eligibility, recruitment and withdrawal of consent

The site-specific research staff are embedded within the LC clinics and will identify potential participants from the clinic list of new referrals. All new referrals will be sent the trial information sheet and contact details for the research team prior to attendance at their first appointment included in the appointment notification letter. This will give participants time to consider taking part prior to their appointment. In instances where the potential participant has not received information then information will be provided at time of approach or clinic visit. The individual will be given as much time as necessary to decide regarding participation and may consent to the nested, adaptive platform randomised drug trial on that day if they wish or return within the 7-10 days post clinic appointment as per patient choice. The individual can contact the team prior to their clinic appointment and ask any questions they may have and, if willing, make an appointment for eligibility assessment and consent prior to clinic, at the time of clinic or in the 7 -10 days after their clinic appointment. Only consent for data collection and blood collection can be taken prior to clinic. Some individuals interested in participating in the trial may have “brain fog” and, therefore, may require more time to consider taking part.

Randomisation into the drug study can only occur after confirmation of eligibility by the clinic team and trial team. Potential participants will also be given information about the study by their treating physician at the LC clinic appointment. Eligibility to the embedded drug trial must be confirmed by the PI or suitably delegated person as per delegation log, following results of clinical assessments, to confirm the participant meets all the inclusion criteria and none of the exclusion criteria. Individuals who are ineligible or opt not to participate in the drug trial will be asked to consent for data collection or data collection plus blood samples for investigation in diagnostic and pathogenesis sub-studies.

The following must be available to confirm eligibility for the nested drug trial:

- Participant full medical history (at LC clinic visit)
- Concomitant medication review (at LC clinic visit)
- Physical Examination (at LC clinic visit)
- Urine pregnancy test within 7 days prior to the first dose of investigational medical product, IMP (in women of childbearing potential)
- Estimated glomerular filtration rate (eGFR) (within last 6 months)
- Liver function test (within last 6 months)
- Full blood count (FBC) (within last 6 months)
- Coagulation screening (within last 6 months)

Site-specific research practitioners embedded in LC Clinics will identify new referrals and, if from within a PCN with Coverscan^™^ as usual care, will arrange a scan appointment 4 weeks prior to first LC clinic appointment (usual care within this PCN, following the same pathway as other clinical appointments). Similarly, individuals referred from PCNs allocated to “Living with COVID recovery^™^” app will be identified and referred to the physiotherapist/s assigned to delivery of this app, who will contact participants directly (as usual care). An appointment with the participant may be required prior to LC clinic to discuss the App, wherever possible or within 7 -10 days of the first LC clinic appointment. We will explain the value of data collection from all participants for the cluster trial and contribution to the blood sample collection, to engage with those participants who may not be eligible for the drug trial but can make a valuable contribution to the study. For individuals where English may not be their first language, the information will contain details of how they can take part assisted by an English-speaking relative/friend.

Participants are free to withdraw at any time from the trial without giving reasons and without prejudicing further treatment and must be provided with a contact point where further information about the trial may be obtained. Data and samples collected up to the point of withdrawal will be used with consent after withdrawal. Any intention to utilise such data shall be included and outlined in the consent form. It will be made clear that, if data has been anonymised and aggregated, it will not be possible to identify individual data for withdrawal. It will also not be possible to identify any one person from such data.

Participants will be notified that their identifiable data (Name, date of birth, NHS Number, and contact details) contact be shared with LCTU to enable contact for follow-up within the study, and for well-being reporting if needed. This trial will only recruit participants who are able to consent. If a participant later becomes incapacitated, the participant shall be withdrawn from the trial and no further data will be collected or any trial investigation or assessments conducted. It is not expected that this would be disease progression within this group. Assessments relating to safety issues arising from participation will continue until the trial ends but will be the remit of the patient’s clinician.

Participants allocated to the cluster with the Living with COVID Recovery^™^ digital app will be informed that data related to their symptoms, physical and mental health, heart rate and physical activities, through the use of the app, will be used to inform their care plan by the Multidisciplinary Team at the LC clinics. The App forms part of their usual care and is part of the clinical record: data from consented individuals will be used by the trial team to assess use of the app (level of engagement) and use of content-specific areas within the app.

### Assessment and Follow-up

All Participants, once consented, regardless of participatory arm, will be invited to complete assessments at baseline, 12 and 24 weeks.

#### Baseline Visit

Demographics, full medical history (from January 2020 when COVID was recognised as circulating in the UK, including comorbidities and all COVID-related illnesses, treatments, and vaccinations); history of concomitant medication including current, pre- and post-COVID-19 and over-the-counter medications; and physical examination (including weight, height, oral temperature, resting pulse, and blood pressure) will be recorded.

Routine clinical blood tests performed in primary care, and as a result of clinic assessment at point of referral, will be collected as part of trial data. Blood tests will differ by clinic, based on differences in usual care and service capacity and capability. Individuals who consent for research blood sampling and for further research will have approximately 60mls blood taken for translational sub-studies and biobanking (**Appendix 3**). All participants who have consented for bloods will be asked to confirm if they would be willing to be contacted at 12 or 24 weeks for further blood sampling.

Standard investigations differ from clinic to clinic. To evaluate usual care, the study will not require a standardised testing strategy; but will collect any, and all data related to individual consented participants tests and investigations (either ordered by the GP at time of referral or ordered as a result of LC clinic appointment). This will include all data related to Coverscan^™^, which will be provided as an uplift to usual care in randomised areas. Examples of functional tests in some clinics are detailed in **Appendix 4**. If the participant consents to the drug trial, a prescription or the IMP will be issued and the first dose taken at the clinic (depending on the location of the recruiting site at the participating centres) or at the time of randomisation or at home for those sites using an IMP delivery service (date of first dose to be confirmed via telephone by site research team) The Patient Reported Outcome Questionnaires and functional tests to be completed are listed in **Appendix 5** and **Table 2**.

#### 12-Week Assessment Visit

Patient Reported outcome measures and activities will be collected at 12 weeks (+ 7 days) to record primary and secondary outcome measures. This may not be possible in all individuals due to their level of fatigue. Participants will be supported by the flexibility of data collection processes to contribute as much as they are able. Patient reported outcome measures will be collected either at the LC clinics, by post, entered by participants themselves via secure electronic link or over the phone as per participant choice. Non-patient reported data will be extracted from the clinical record by the site-specific research staff and entered in the trial database (listed in **Appendix 6**).

#### 24-Week Assessment Visit

All participants will be followed up for 24 Weeks (+ 7 days) from enrolment date by LCTU staff. Participants will be given the choice of completing questionnaires online using an individualised secure link, by postal return or via telephone for participants requiring support. In the event of receiving an incomplete outcome assessment, participants will be contacted by phone within 7 days of the team receiving the form (e-form or paper) to complete missing information with the individual. Data will be entered into the trial database by a LCTU staff. Patient Reported Outcome measures and functional assessments recorded are listed in **Appendix 7**.

### Outcomes

#### Primary Outcome

Fatigue is the dominant symptom in 60% of patients and LC clinics are being commissioned to provide rehabilitation for 12 weeks or less, therefore, an outcome at 12 weeks is a pragmatic primary outcome measure. Fatigue Assessment Scale (FAS) is a 10-item, validated questionnaire in individuals with chronic diseases, including LC(21)(22). Therefore, it is a pragmatic primary end point for the STIMULATE-ICP trial. Five questions reflect physical fatigue and five questions (questions 3 and 6-9) reflect mental fatigue. The total score ranges from 10 to 50.

#### Secondary Outcomes

Secondary outcomes include:

1. FAS at 24 weeks
2. Health related quality of life (EQ-5D-5L)(23)
3. Mental health (GAD-7)(24)
4. Depression (PHQ-9)(25)
5. Medical Research Council Dyspnoea Score(26)
6. Perceived Deficit Questionnaire (PDQ-5)(27)
7. Work and Social Adjustment Scale (WSAS) [Question 4 from Productivity Cost Questionnaire (iPCQ) for absenteeism and Question 8 from iPCQ for presenteeism added](28)(29)
8. Short Form Questionnaire (SF-12)(30)
9. Cognitive Failure Questionnaire (CFQ) if an individual scores 3 or more on PDQ5 (individuals receive an email to complete this questionnaire online via a secure password and patient ID number)(31,32)
10. Organ impairment and healthcare utilisation
11. Cost-effectiveness of ICP (a separate health economic evaluation is planned(18) and the protocol will be published separately)
12. Process outcomes for different ICP components
13. Blood investigations (e.g., genomic, proteomic, metabolomic/ lipidomic, functional T cell and live virus neutralisation assays and endocrine analyses)
14. Linked electronic health record data to monitor healthcare utilisation and outcomes.

#### Exploratory Outcome Measures / Endpoints

There is ongoing research developing a core outcome data set for clinical and research use in individuals with LC. For example, based on a NIHR-funded research programme (Post COVID Core Outcome Set, PC-COS(33)) which conducted an extensive, international Delphi process in over 1500 patients and health professionals internationally, we included consensus physiological/clinical, life impact and recovery domains. As our trial progresses, we will collaborate with PC-COS in their work to iteratively and pragmatically develop LC core outcomes.

### Data Collection

All e-(electronic) and p-(paper) clinical research forms (CRFs) must be completed and signed by designated, authorised research team staff. The PI is responsible for accuracy of all pCRF data. Information from pCRFs will be entered into an eCRF within the electronic trial database by the site team directly for the baseline and 12 -week assessment (where participants attend in person). For virtual clinics and where the patients are not returning to the clinic in person, participants will be given the option of completion via postal, e-link (directly into database for self-completion) or over the telephone for participants requiring support. All the CRFs will be entered into the trial database within 15 days of data received by the site staff or CTU staff as applicable. **Table 2** summarises objectives and outcome measures and when they will be collected, and the means of data collection are detailed in the previous section. All data are managed (further details in **Appendix 8**) in accordance with LCTU standard operating procedures (DM-03 and DM-05) and a trial-specific Data Management Plan will be in place before opening to data collection, including details of database (software, design and lock) and data (validation plan, entry, quality checks, queries and security). Where data are transferred electronically this will be in accordance with the UK Data Protection Act 2018 as well as UCL and LCTU Information Security Policy and Trust Information Governance Policy.

Individuals referred from a PCN who are allocated to receive the Coverscan^™^, will be sent an appointment for the scan 4 weeks prior to their LC. The scan results will be assessed and reported on by a specialist radiologist group supported by Perspectum. Reports will be returned to clinic site and become part of the clinical record. Reports should be available to clinicians at the time of the clinic visit to inform decision-making in clinical care. Uptake of Coverscan™ will be entered into the trial database for analysis.

Routine standard of care clinical data from patients undergoing investigations at participating LC clinics will be captured through patient-facing questionnaire pCRF (paper clinical research form) and research staff completed pCRF (clinical data and investigations collected from the clinical record and physical assessment) by a member of the trial team at the site.

Participants will be consented for use of NHS data regarding medical history (including hospital and outpatient attendances, prescriptions, and other relevant information) from January 2020 and up to 1 year following enrolment in the study. Data capture will be via NHS Digital using NHS number.

The CI, Sponsor and trial statisticians will have access to the full pseudonymised dataset prior to database lock and data analysis at the end of the trial. Collaborators and the PIs have access to anonymised data extracts at the end of the study

### Analysis

The trial will be analysed and reported using “Consolidated Standard of Reporting TrialsD (“CONSORT”)(34) and International Conference on Harmonisation E9 guidelines(35). A CONSORT diagram will report flow of participants in all arms of the cluster trial. Baseline characteristics of all consenting participants will be reported by frequency and percentage for categorical variables, and for continuous variables by mean and standard deviation (or median and inter-quartile range for non-normally distributed data).

#### Primary and secondary outcomes

Primary analysis of the trial will be a complete case analysis, carried out using all available outcomes, according to the cluster randomised allocation of the participant’s PCN (regardless of treatment pathways received) or the stratified randomisation of the individual participant (regardless of treatment received). The primary outcome, FAS at 12 Weeks, is expected to be approximately normally distributed(36). A multi-level model analysis will be used to evaluate effects of Coverscan^™^, digitally enabled community rehabilitation, and their interaction, on FAS at 12 weeks adjusting for baseline FAS, with clinic and PCN as random effects.

The principal analysis population for these estimates of the effects of Coverscan^™^, digitally enabled community rehabilitation, and their interaction, will be those participants (up to 1130) who consented to data collection for the cluster RCT and were allocated to usual care (either by randomisation to no drug in the drug trial or being willing for data collection, but not eligible for the drug trial, or unwilling to be randomised into the drug trial). This analysis will provide the most straightforward answer to the primary objective in the broadest group of participants. A similar and more pragmatic multi-level model analysis of the effects of Coverscan^™^, digitally enabled community rehabilitation, and their interaction, will be applied to all participants who consent to data collection in the cluster trial regardless of participation in the drug trial. This analysis will provide an answer to the primary objective in the presence of other treatments including drug treatments and will allow comparison to results from the principal analysis set.

The principal analysis population for drug vs. usual care (no drug) comparisons within the drug trial will be the participants randomised to the control treatment pathway, i.e. no Coverscan + usual care rehabilitation. Participants randomly allocated to each drug arm will be compared to those randomly allocated to usual care while both that drug and usual care are options for randomisation (concurrent usual care controls). A multi-level model for FAS, adjusting for baseline, with gender as a fixed effect and clinic and PCN as random effects, will be applied to the drug trial data. By restricting the comparisons of the effects of the drugs vs. usual care on fatigue to those participants allocated to the treatment pathway incorporating the lowest level of intervention, and potentially the least variation in care, this will give the cleanest and least biased comparison. The total population of participants who are randomised into the drug trial will be used to study the effects of the drugs in combination/interaction with the pathways. A multi-level model for FAS, adjusting for baseline, with gender as a fixed effect and clinic and PCN as random effects, will be applied to data from all participants of both the cluster trial and the drug trial to evaluate effects of Coverscan^™^, digitally enabled community rehabilitation, and their interaction, in combination with the participants’ allocated drug treatment. For all drug treatments, particularly any introduced after the start of the trial, comparisons with participants randomised to no drug will include only those participants who could have been randomised to the relevant drug treatment at the time of their randomisation. This will reduce the potential for bias and have the effect of ensuring that only concurrent participants allocated to no drug are compared to participants randomised to drug treatments.

Secondary outcomes on an interval scale will be analysed similarly to the primary outcome, adjusting for baseline outcome measure, clinic and PCN as a random effect. The principal analysis set for estimates of effects of Coverscan^™^, digitally enabled community rehabilitation, and their interaction, on these secondary outcomes (EQ-5D-5L, GAD-7, PHQ-9, WSAS, PDQ-5, CFQ, SF-12, MRC Dyspnoea score) being participants who consented to data collection for the cluster RCT and were allocated to usual care (either by randomisation to no drug in the drug trial or being willing for data collection, but not randomisation into the drug trial). Where appropriate, alternative methods will be applied to outcomes with skewed distributions. Secondary outcomes on an interval scale will be compared between drug treatment arms using the same methods as for the primary outcome, using a multi-level model for the relevant outcome, adjusting for baseline, with gender as a fixed effect and clinic and PCN as random effects, within the control treatment pathway.

Patterns of missingness will be summarised. In modelling, complete case analysis will be the main analysis. For missing outcome data, we will use a threshold approach as a sensitivity analysis. Imputation based on thresholds will be implemented such that missing values will be replaced by values from the lower and upper quantiles of the distribution, e.g. 90th quantile, 75th quartile, 25th quartile, 10th quantile, and minimum. Other patient reported secondary outcomes with missing items will be addressed using the recommended methods for missing items for that scale.

#### Sensitivity and subgroup analyses

A sensitivity analysis of the effects of Coverscan^™^, digitally enabled community rehabilitation, and their interaction, will include only those who are randomly allocated to receive usual care (no drug) in the drug trial. Results will be compared to those of the (broader) principal analysis population which also includes those unwilling or unable to be included in the drug trial.

Health outcomes and health utilisation data taken from medical records at 12 months will be described using summary statistics for all participants and for subgroups defined by sub-phenotype (clinical or pathophysiological) at baseline. Analyses of all subgroups will be considered as exploratory. Subgroups may include cardiorespiratory, neuropsychiatric and “mast-cell activation type” (including rashes, joint pain and gastrointestinal disturbances and serositis symptoms). These subgroups are not mutually exclusive as some participants may belong to more than one subgroup depending on their symptoms.

If the Living with COVID Recovery^™^ application is found to be beneficial, further exploratory analyses will explore factors associated with greater benefit. Some participants allocated to this application will not be able to access it as they may lack a suitable mobile phone or tablet. Other participants will have suitable access but may choose to interact with it on a more or less extensive basis. Amount of access will be available as a patient reported variable and as summary variable from within the application.

#### Interim Analysis

The first interim analysis point will be after 1200 participants are recruited to the cluster randomised trial and the remaining interim points will be after every 600 participants recruited. The interim analysis time points will apply to both the cluster and drug trials.

Interim analysis of the cluster interventions will take place among the participants receiving usual care only (by allocation in the drug trial to no drug or by refusing or being ineligible for the drug trial). A pathway (a combination of the two cluster interventions) could be found to be superior after the first 1200 participants and every 600 participants thereafter in the study. A statistically significant p-value at the 0.001 two-sided level is required for a pathway to be deemed superior at interim analysis. This is to account for multiple testing (Bonferroni type correction). The analysis at 4520 individuals in total will be at the conventional 0.05 two-sided level. There are practical and operational reasons why a single pathway cannot graduate and become the standard pathway within the recruitment period of this trial.

For a drug to graduate early and become part of the backbone treatment of the trial, its two-sided p-value at each of the interim time points will need to be <0.001. The final principal analysis for each drug, including only those participants in the standard of care cluster pathway, will be with at least 200 participants in the active arm and 200 concurrent controls, and will use the conventional 2-sided 0.05 level. This analysis has 85% power if Cohen’s D is 0.3 and 97% power if Cohen’s D is 0.4.

### Power Calculation

#### Main study

The cluster randomised study takes the form of a 2×2 factorial trial where the unit of randomisation is the PCN. The ze calculation was based on the power to detect an interaction effect of clinical importance of 3 points (40) on the FAS scale (41) rather than on the size of the main effects themselves. Therefore, the sizes of the main effects are not specified below. For context, the difference in means of FAS between people who say they have or have not recovered from LC was 9 points in a recent observational study(41).

The sample size was calculated using PASS v21.0.1(2021) for a 2×2 cluster factorial trial. The study was powered to detect an interaction on the FAS scale between Coverscan^™^ and Living with COVID Recovery^™^. The sample size does not depend on the size of main effects of both individual interventions. Based on published data the standard deviation on the FAS scale for individuals with LC is estimated at 6 units. An interaction effect of 3 points (40) on the FAS can be detected with just over 90% power and (two-sided) significance level of 0.05 with 960 participants in 48 PCN clusters of 20 participants, assuming a conservative intra-cluster correlation coefficient (ICC) of 0.02. If there is dropout of 15% (to be conservative) and assuming missingness is roughly equal across the arms, then the number of participants must be inflated by a factor of 100/85 to 1130 (approximately 56.5 centres of 20 participants).

Experimental estimates of the prevalence of symptoms that remain 12-weeks after COVID infection range from 3.0% based on tracking specific symptoms, to 11.7% based on self-classification of LC, using data to 1 August 2021 (3). One LC clinic has ∼25 new individuals per week(16), suggesting there might be around 1300 new individuals annually per clinic. Most are expected to give consent to data collection even if not willing to consent to randomisation into a drug study. LC referral rates range from 0-2.5/1000 per PCN. Assuming this rate will not change over the foreseeable future, this will ensure that our 6-10 LC clinics will provide sufficient patients for this study. Moreover, there is a backlog of people waiting for their first LC clinic appointment (from private communication).

#### Nested Drug Trial

The total number of individual drugs which will be tested on the platform is currently unknown. The total sample size required in an adaptive platform drug trial, with uncertainty in the timing of the introduction of further drugs, cannot be set in advance, but would need to be updated in consultation with the IDMC and funder throughout the trial at set interim points in data collection.

The treatment effect is unknown, at this time, for the drugs. The plan is to perform interim analysis after 1200 participants in total are recruited and after every 600 participants thereafter. With 200 participants per active arm and corresponding concurrent controls the study has 85% power to detect a Cohen’s D of 0.3 on FAS (difference between means in two treatment arms divided by the standard deviation of the data) at the 0.05 two-sided alpha level. If this number is increased to 300 for the same effect size the power is 95%.

#### Planned Recruitment Rate

To plan and cost the trial a nominal maximum of 4520 recruited participants was proposed based on up to 4 (drug and usual care) parallel arms of up to 1130 evaluable participants. The nested drug trial aims to recruit up to 4520 individuals from a possible pool of 30 000 individuals from 6-10 PCN areas of England. The trial will continue to recruit participants at least until the recruitment target of 1130 participants for each of the initial arms of the adaptive trial has been reached. If the adaptive trial requires the full 4520 participants, it is estimated it will take approximately 10 to 12 months for the recruitment to complete and by the 16^th^ month from the start of the trial, the last visit of the last patient would be completed in the third quarter of 2023. The projected recruitment rate, when all clinics are open to recruitment, is 600 participants per month.

### End of trial

The study is deemed to have ended following the last data collection point within the trial. This will be 12 months following enrolment of the last participant in the trial. Thereafter, there will be a 6-month period for data analysis and reporting. The chief investigator (CI) and/or trial steering committee (TSC) have the right at any time to terminate the trial for clinical or administrative reasons. The funder may withdraw funding in the event of futility, study misconduct, or other unanticipated events. In such instances, the Sponsor will notify the MHRA within 15 days. The end of the trial will be reported to the REC and Regulatory Authority within the required timeframe if the trial is terminated prematurely. Investigators will inform patients of any premature termination of the trial and ensure that the appropriate follow up is arranged for all involved. Following the end of the trial a summary report of the trial will be provided to the REC and Regulatory Authority within the required timeframe. The Sponsor will notify MHRA at the end of the clinical trial within 90 days of its completion.

The trial may be stopped before completion on the recommendation of the TSC, IDMC (independent data monitoring committee) or the Sponsor and CI. All safety data will be reviewed and a decision on continuation will be made by the IDMC with input from the Sponsor. A single arm of the drug study may be stopped by the IDMC on grounds of harm at each stopping point. Additionally, the IDMC may recommend a single arm of the drug study be halted in the event of the death of a single participant, directly attributable to the study drug.

At the end of the trial, all essential documentation and the trial dataset will be prepared for archiving and transfer to the Sponsor organisation by Lancashire CTU. Participating sites will be asked to archive for a minimum of 25 years from the declaration of end of trial. Essential documents are those which enable both the conduct of the trial and the quality of the data produced to be evaluated and show whether the site complied with the principles of Good Clinical Practice and all applicable regulatory requirements. The Sponsor will notify sites when trial documentation can be archived. All archived documents must continue to be available for inspection by appropriate authorities upon request.

### Oversight

The Trial Management Group (TMG) is responsible for the management of STIMULATE-ICP and is led by AB (CI) and MH (lead clinical principal investigator, PI). In addition, TMG comprises trial investigators and relevant staff from LCTU and PPI members, who meet regularly to discuss the progress of the trial. LCTU is responsible for day-to-day trial management, is the data custodian, and will conduct central and on-site monitoring of sites and data. STIMULATE-ICP is managed according to Good Clinical Practice guidelines. LCTU will act to preserve patient confidentiality and prevent disclosure or reproduction of identifiable patient information by encryption to ensure anonymity. Procedures for handling, processing, storing and destroying data are compliant with the Data Protection Act 1998. The TSC and IDMC have been convened as trial oversight committees. The TSC is independently chaired by Professor Patrick Mallon and includes clinicians, academics, and PPI representatives, along with CI and lead clinical PI. TSC monitors trial progress. The IDMC is chaired by Professor Peter Langhorne, includes both clinical and statistical expertise, and advises the TSC, independent of the trial team, sponsor and TSC, making recommendations on continuation, or not, of the trial. Membership of each committee depends on accepting terms of reference and declaration of conflict of interest. Any major protocol modifications (such as changes to eligibility criteria, outcomes, analyses) will be communicated to trial registry (ISRCTN), research team, REC and HRA and the public via the study website.

## Discussion

In individuals with non-hospitalised LC, the STIMULATE-ICP trial represents the first major, pragmatic trial to study (i) drugs in a nested platform design, (ii) components of an ICP from pre- to post-clinic care, and (iii) fatigue as a primary endpoint. Given the national and international burden of LC, STIMULATE-ICP is important to investigate feasible, generalisable and scalable improvements in care through: (i) widely-used, re-purposable drugs, (ii) accessible interventions (Coverscan™ and Living with COVID Recovery™), and (iii) study sites varying in geography and clinical service. This paper presents the protocol (Version 2.1, 05/05/2022) and the full protocol is available on the NIHR and study (https://www.stimulate-icp.org/) websites. The SPIRIT checklist(37) for the trial protocol is included in **Appendix 9**.

Recruitment started in July 2022. At the time of first manuscript submission, data collection for the trial was ongoing and due to be complete in July 2023. The full trial results will be disseminated in Q3 2023 through presentations at national and international conferences, publication in peer reviewed medical journals and regular interaction with media (mainstream and social). As well as the non-trial aspects of STIMULATE-ICP (described in the methods), there are now several nationally funded observational research programmes are underway, including LOCOMOTION, TLC and PC-COS studies(17). In addition, there are now several ongoing clinical trials in LC:

### Phase 1/2

RSLV-132 (https://clinicaltrials.gov/ct2/show/NCT04944121)

Zofin (https://clinicaltrials.gov/ct2/show/NCT05228899)

Axcella-1125 (https://www.clinicaltrials.gov/ct2/show/NCT05152849)

### Phase 3

HEAL-COVID (https://heal-covid.net/)

However, to our knowledge, ongoing studies are neither platform studies nor specifically focused on individuals with non-hospitalised LC.

STIMULATE-ICP will provide up-to-date information about investigation and rehabilitation components of the clinical pathway for individuals with LC, as well as developing the evidence for therapeutics in this patient population.

#### Data Sharing

The protocol does not report results and no further data are available. Deidentified research data will be made publicly available when the study is completed and published.

## Acknowledgements

STIMULATE-ICP team:

<u>UCL:</u> Amitava Banerjee, Paula Lorgelly (Auckland), Elizabeth Murray, Hakim-Moulay Dehbi, Hugh Montgomery, Sarah Clegg, Henry Goodfellow, Mel Ramasawmy

Yi Mu, Sampath Weerakkody, Ileana Selejan, David Sunkersing, Ashkan Dashtban

<u>UCLan</u>: Caroline Leigh-Watkins, Denise Forshaw, Gordon Prescott

<u>RCGP:</u> Gail Allsopp

<u>University of Liverpool:</u> Mark Gabbay, Gregory Lip, Dan Cuthbertson, Dan Wootton, Nefyn Williams

<u>University of Hull:</u> Mike Crooks

<u>Hull University Teaching Hospitals Trust:</u> Angela Green

<u>University of York:</u> Christina Feltz van der Feltz-Cornelis, Jenny Sweetman, Han-I Wang, Natalie Smith

<u>University of Leicester:</u> Kamlesh Khunti, Lauren O’Mahoney, Rachael Evans

<u>University of Exeter:</u> David Strain

<u>Royal Devon University Healthcare NHS Foundation Trust:</u> Rachel Botell

<u>University of Leicester:</u> Kamlesh Khunti, Lauren O’Mahoney, Rachael Evans

<u>University of Southampton:</u> Nisreen Alwan, Donna Clutterbuck

<u>University of Sussex:</u> Marija Pantelic

<u>Living with Covid Recovery:</u> Chris Robson

<u>Perspectum:</u> Mike Brady, Rajarshi Banerjee, Cat Kelly, Angela Barone, Johannes Alberts, Rob Suriano

<u>PPI:</u> Lyth Hishmeh, Emily Attree, Jasmine Hayer, Rita Mallinson Cookson, Rachel Hext, Andrew and Rachel Williams, Mag Leahy, Antony Loveless, Clare Loveless, Kim Horstmanshof

Collaborators:

<u>Royal College of Speech and Language Therapists:</u> Gemma Clunie

<u>NNEdPro Global Centre for Nutrition and Health:</u> Dominic Crocombe, Shane McAuliffe

Independent Data Monitoring Committee: Peter Langhorne (chair), Chris Sutton

Trial Steering Committee: Independent members: Patrick Mallon (chair), Marion Mafham, Ondine Sherwood (PPI). Other members: Matthew Sydes

UCL/UCLH Joint Research Office (sponsor): Michelle Quaye, Farhat Gilani, Yusuf Jaami, Nikki Cleary, Judy Jones, Robin Holas

## Contributors

Design: AB, ECW, MH, DF and CW led the trial design. Writing: DF and AB wrote the first draft of the manuscript and all authors contributed to the final manuscript. Statistics: GP and HMD led the statistical planning and analysis. PPI: LH and EA led the PPI team.

## Funding

This trial is part of a National Institute for Health Research (NIHR: COV-LT2-0043)-funded programme (**Figure 2**)(17), with epidemiologic and mixed methods studies(18), including care inequalities and transferability to other LTCs (IRAS 303958) and this complex intervention trial (IRAS 1004698). Funding for the transport, storage, processing and biobanking of blood and MRI is supported by Perspectum Ltd. The funders had and will not have a role in study design, data collection and analysis, decision to publish, or preparation of the manuscript. The views and opinions expressed therein are those of the authors and do not necessarily reflect those of the NIHR, NHS, the Department of Health and Social Care, or the sponsor.

## Patient consent

Full, informed individual consent for all aspects of the trial.

## Competing interests

WDS holds research grants from Bayer, Novo Nordisk and Novartis and has received speaker honoraria from AstraZeneca, Bayer, Bristol-Myers Squibb, Merck, Napp, Novartis, Novo Nordisk and Takeda, outside the submitted work. WDS is supported by the NIHR Exeter Clinical Research Facility and the NIHR Collaboration for Leadership in Applied Health Research and Care (CLAHRC) for the South West Peninsula. MGC has received honoraria, fees for advisory boards, and non-financial support from AstraZeneca, BI, Chiesi, GSK, Novartis, and Pfizer and grants from AstraZeneca, BI, Chiesi, and Pfizer. CR is director of Living With, which developed Living With COVID Recovery™ and other digital health interventions used in healthcare systems including the NHS. RB is CEO of Perspectum, which developed Coverscan™. AB has received research funding from NIHR, British Medical Association, Astra Zeneca, National Institute of Aging and European Union. AB is a Trustee of Long COVID SoS. All other authors report no competing interests.

## Ethical approval

The Protocol (version 2.1; 5/5/2022) and other trial-related documentation (and any amendments) received favourable ethical opinion from NRES Committee South Central - Berkshire Research Ethics Committee (reference: 21/SC/0416). All participating sites obtained local approvals prior to patient recruitment. All participants will give full, informed consent to participate in the trial.

## Provenance and peer review

Not commissioned; peer reviewed for ethical and funding approval prior to submission.

## Open Access

This is an Open Access article distributed in accordance with the terms of the Creative Commons Attribution (CC BY 4.0) license, which permits others to distribute, remix, adapt and build upon this work, for commercial use, provided the original work is properly cited. See: http://creativecommons.org/licenses/by/4.0/

## Notes

### Competing Interest Statement

I have read the journal's policy and the authors of this manuscript have the following competing interests: WDS holds research grants from Bayer, Novo Nordisk and Novartis and has received speaker honoraria from AstraZeneca, Bayer, Bristol-Myers Squibb, Merck, Napp, Novartis, Novo Nordisk and Takeda, outside the submitted work. WDS is supported by the NIHR Exeter Clinical Research Facility and the NIHR Collaboration for Leadership in Applied Health Research and Care (CLAHRC) for the South West Peninsula. MGC has received honoraria, fees for advisory boards, and non-financial support from AstraZeneca, BI, Chiesi, GSK, Novartis, and Pfizer and grants from AstraZeneca, BI, Chiesi, and Pfizer. CR is director of Living With, which developed Living With COVID RecoveryTM and other digital health interventions used in healthcare systems including the NHS. RB is CEO of Perspectum, which developed CoverscanTM. AB has received research funding from NIHR, British Medical Association, Astra Zeneca, National Institute of Aging and European Union. AB is a Trustee of Long COVID SoS. All other authors report no competing interests.

### Clinical Trial

ISRCTN 10665760

### Funding Statement

This trial is part of a National Institute for Health Research-funded programme (NIHR: COV-LT2-0043). The CI is AB. Funding for the transport, storage, processing and biobanking of blood and MRI is supported by Perspectum Ltd (CI-AB). The funders had and will not have a role in study design, data collection and analysis, decision to publish, or preparation of the manuscript.

### Author Declarations

South Central - Berkshire Research Ethics Committee (reference: 21/SC/0416) of the Health Research Authority UK gave ethical approval for this work.

### Summary of Updates

We have added the supplementary file with Appendices which was omitted from the prior version

