## Appendix 1 for "STIMULATE-ICP: A pragmatic, multi-centre, cluster randomised trial of an integrated care pathway with a nested, Phase III, open label, adaptive platform randomised drug trial in individuals with Long COVID: a structured protocol"

**Supplementary materials**

| Appendix 1:  Rationale for initial drugs in drug platform | 2 |
| --- | --- |
| Appendix 2:  Additional Inclusion Criteria for the nested, platform randomised drug trial | 5 |
| Appendix 3:  Samples for Future Research and Biobanking Storage | 7 |
| Appendix 4:  Functional tests in some Long Covid clinics | 8 |
| Appendix 5:  Functional and Patient Reported Outcome Assessments at Baseline | 9 |
| Appendix 6:  12-Week Assessment Visit | 10 |
| Appendix 7:  24-Week Assessment Visit | 11 |
| Appendix 8:  Data management plan | 12 |
| Appendix 9:  SPIRIT 2013 Checklist | 13 |

***Appendix 1:***

***Rationale for initial drugs in drug platform***

### Loratadine and Famotidine (H_1_ + H_2_ Receptor Blockade)

Loratadine and Famotidine are both histamine receptor antagonists. In combination they inhibit both the H_1_ and H_2_ receptors. Famotidine is an effective competitive H_2_ receptor antagonist. It reduces the concentration and amount of acid and pepsin of the gastric juices. The effect of oral administration is rapid, long lasting when used at the recommended dosage and it is effective with relatively low concentration in the blood. The duration of its effect, plasma concentration and secretion in the urine are dose-dependent. Famotidine is licensed as an over the counter (OTC) treatment for dyspepsia/gastric ulceration. Loratadine is also an OTC treatment licensed for treatment of mild allergic symptoms including seasonal rhinitis.

Individuals with Long COVID (LC) are hypothesised to have a persistent inflammatory process that may drive the symptoms of fatigue and myalgia. In a small minority of individuals, more specific symptoms suggestive of mast-cell activation, including rashes, diarrhoea and flushing may be present. Histamine receptor antagonists are commonly used to treat mast-cell activation in other conditions.

Mixed reports of efficacy of histamine receptor antagonists in acute COVID-19, particularly reduction in oxygen requirements and possibly mortality, have led to ongoing clinical trials of these treatments in acute COVID-19 (27). The suggested mechanisms of action in acute COVID-19 are reduction in inflammation due to suppression of T-cell mediated cytokine release, however, available data are very limited. In an observational cohort study, 49 patients with LC were compared to matched asymptomatic fully recovered COVID-19 individuals without LC. Individuals with LC were offered treatment with combined histamine H1/H2 blockade, using H1 (Loratadine 10 mg once daily or Fexofenadine 180 mg twice daily (not being used in this study) and H_2_ (Famotidine 40 mg once daily or Nizatidine 300 mg once daily (not being used in this study)) for a minimum of 4 weeks (28), based on data from acute COVID-19 suggesting histamine receptor antagonist therapy improved symptoms (29, 30). In this preliminary observational study, individuals with physician-diagnosed LC reported a 48% reduction in symptom burden after 4 weeks of combined histamine receptor antagonist therapy, compared to baseline. When compared to recovered non-hospitalised individuals, individuals with LC had significantly lower circulating CD4 but not CD8 effector memory cells, suggesting that this T cell subset may be involved in LC pathology (28). These data, however, have significant limitations, and the findings require testing in a formal clinical trial.

In this study we will be using famotidine 40mg once daily in combination with loratadine 10mg OD, ensuring combined histamine receptor antagonist activity across the H_1_ and H_2_ receptors. Both drugs are being used within their licensed dose and are safe to be co-administered.

### Colchicine

Colchicine inhibits cellular transport and mitosis by binding to tubulin and preventing its polymerisation as part of the cytoskeleton transport system. Colchicine has a short half-life of 9-12 hours and is prescribed as a BD (twice daily) dosing regimen. Standard doses for acute gout range from 500mcg BD to 2mg BD, depending on the dose response. Colchicine has a wide range of anti-inflammatory effects, including inhibition of certain inflammasomes (cytosolic pattern recognition receptor systems that are activated in response to detection of pathogens in the cytosol) (31), (32). Evidence shows that inflammasomes are activated in COVID-19, and the degree of activation is correlated with disease severity ( (33)).

Colchicine has been shown to have cardiovascular benefit in individuals with coronary artery disease and pericarditis at a dose of 500mcg BD. Its primary mechanism of action is reduction of serositis, inflammation of membranes around joints and viscera. Individuals with LC frequently complain of symptoms suggestive of serositis, either atypical chest pain that may indicate pericarditis, costochondritis or pleural inflammation, or joint pain in the absence of clinically evident inflammation. Disease severity in LC correlates with myocardial damage on Coverscan™ (15). Of the non-hospitalised individuals with LC at UCLH, 20% report chest pain and or palpitations, 60% report shortness of breath. Standard investigations are frequently normal: prolonged rhythm monitoring typically shows resting sinus tachycardia, chest pain is generally atypical with predominantly normal echocardiogram, Troponin T and ECG. In a prospective cohort of over 50 individuals (median age 43, 69% female) reporting persistent chest pain underwent cardiac MRI (CMR). 26% had evidence of myocarditis-pattern late gadolinium enhancement and/or evidence of abnormalities, (meeting diagnostic criteria for myocarditis (34). T1/T2 abnormalities were seen in 29%. Individuals with LC with myocarditis contrasted to individuals admitted to hospital with moderate/severe respiratory COVID (mean age 64, 66% male). However, the prevalence of myocarditis is strikingly similar (35). To date, we have treated in excess of 200 LC individuals with abnormal CMR empirically with colchicine 500mcg BD, 9/10 reported significant symptomatic improvement within 3 months.

### Rivaroxaban (Low Dose Anticoagulation)

Rivaroxaban is an oral factor Xa inhibitor, inhibiting the clotting cascade. Individuals with LC who complain of marked exertional fatigue with abnormalities on the 6-minute walk test, have evidence of microvascular anaerobic respiration, despite normal oxygenation in peripheral blood. This suggests that while adequate haemoglobin-bound oxygen is present in blood, the dissociated oxygen is unavailable to large muscles on increased aerobic demand during exercise. Abnormalities of the clotting cascade are a marked feature of acute Covid-19, including microvascular thrombi (36). Thrombi in acute Covid-19 are strongly associated with abnormally elevated von-Willebrand Factor (VWF): ag/ADAMTS 13 ratio (37). Recent data are showing longer term risk of VTE with LC. We extended the measurement of VWF Ag/ADAMTS 13 levels to the UCLH community LC cohort (Scully and Heightman, 2021. Unpublished data.) Of 272 patients in LC clinic describing extreme lethargy, headaches and poor exercise tolerance, 81/272 (30%) had an abnormal VWF Ag/ADAMTS 13 ratio of >1.5. Elevated VWF Ag/ADAMTS 13 ratio strongly associated with impaired exercise capacity on a 6-minute walk test: 1.5 compared to 1.1 in patients with normal exercise capacity (p<0.001).

A further 28 patients had blood analysed by flow chamber assay, which measures VWF, platelet binding and in-vitro thrombus formation in real time. 8/28 (29%) had clot formation by 5 minutes. In a small feasibility project, 5 patients were initiated on low dose aspirin (LDA) 75mg daily, selected based on symptom severity and elevated VWF(Ag):ADAMTS13 ratio. In 3/5 patients, improvement in symptoms and VWF(Ag):ADAMTS13 ratio was observed. The remaining two stopped aspirin due to upcoming procedure and bruising. Patients B and C were analysed on the flow chamber pre- and post-aspirin usage with marked improvement in surface coverage from 100% to 29% and 9% respectively on LDA, reporting corresponding symptom improvement.

Evidence of microvascular thrombi in some patients has led to the hypothesis that that endothelial dysfunction due to multiple microvascular thrombi in large muscles may significantly contribute to reduced aerobic capacity and symptoms of fatigue. Low-dose anticoagulation is a safe approach to test this hypothesis, by measuring fatigue as the primary outcome, we will determine if this approach indicates the presence of microvascular thrombi in LC patients. Aspirin was poorly tolerated when tested in our pilot study due to gastro-intestinal side effects. We have included rivaroxaban 10mg once daily in the STIMULATE-ICP trial instead of aspirin due to improved safety profile, easy comparability with other clinical trials using factor Xa inhibitors such as apixaban (HEAL-COVID) and proven efficacy data on prophylaxis of intravascular thrombi. The rivaroxaban regime of 10mg OD proposed in STIMULATE-ICP is a prophylaxis dose, which aims to effectively prevent generation of further microvascular thrombi, while minimising bleeding side effects. This prophylaxis dose is therapeutically equivalent to low-dose aspirin tested in our preliminary cohort.

***Appendix 2:***

**Additional Inclusion Criteria for the nested, platform randomised drug trial**

*N.B: Potential participants with drug-specific contraindications for any arm, including interactions of pre-prescribed essential medication will be consented for data collection but will be excluded from the drug study.*

1. Females of childbearing potential (see definition below) must be willing to use at least an acceptable effective method of contraception during the treatment with investigational medical product (IMP) and for a further 30 days after the last dose. (34).

Such methods include:

a. combined (oestrogen and progestogen containing) hormonal contraception:

I. oral
II. intravaginal
III. transdermal

b. progestogen-only hormonal contraception

I. oral
II. injectable
III. implantable

c. intrauterine device (IUD)

d. intrauterine hormone-releasing system (IUS)

e. bilateral tubal occlusion

f. vasectomised partner

g. male or female condom with spermicide

h. cap, diaphragm or sponge with spermicide

i. sexual abstinence; only true abstinence is acceptable i.e. when this is in line with the preferred and usual lifestyle of the participant). (Periodic abstinence, declaration of abstinence during exposure to IMP and withdrawal are not accepted methods of contraception).

Definition of females of childbearing potential:

For the purpose of this trial, a female is considered of childbearing potential i.e. fertile following menarche and until becoming post-menopausal unless permanently sterile. Permanent sterilisation methods include hysterectomy, bilateral salpingectomy, and bilateral oophorectomy.

A post-menopausal state is defined as no menses for 12 months without alternative medical cause.

2. Male Participants must be willing to use condom during IMP treatment to protect their female partner becoming pregnant and for a further 90 days after the last dose.

3. Patients on pre-existing treatments for the same drug classes MUST undergo a 7-day washout period before being randomised.

*(Patients will be assessed, and if safe to do so, exclude that medication for 7 days, asked if they would be willing to undergo a washout period of at least 7 days before being randomised.)*

*Exclusion Criteria for ALL Participants*

1. Previously hospitalised for COVID-19 infection.

2. Previously referred to a LC clinic.

*Exclusion criteria for nested, adaptive randomised drug trial*

3. Females who are pregnant, planning pregnancy or breastfeeding

4. Known hypersensitivity to any of the study drugs or their excipients

5. Currently taking any of the following drugs:

Probenecid, Sucrafate, Isocarboxazid, Phenylzine, Tranylcypromine of any other CNS depressant (such as diphenhydramine, dextromethorphan, or pseudoephedrine) *(Contraindications to famotidine/loratadine)*

Amiodarone, Aprepitant, Atanazavir, Atorvostatin, Azithromycin, Bezafibrate, Ciclosporin, Ciprofibrate, Clarithromycin, Cobicistat, Croztibib, Darunavir, Diltiazem, Dronedarone, Eliglustat, Erythromycin, Fenobibrate, Fluconazole, Fluvastatin, Fosamprenavir, Gemfibrozil, Idelalisib, Imatibib, Isavuconazole, Itraconazole, Ketoconazole, Letermovir, Lopinavir, Netupitant, Nilotinib, Posaconazole, Pravastatin, Ranolazine, Ritonavir, Rosuvastatin, Simvastatin, Tipranavir, Velpatasvir, Vemurafenib, Venetoclax, Verapamil, Voriconazole *(Contraindications to colchicine)*

Acalabrutinib, Aceclofenac, Acenocoumarol, Alprostadil, Alteplase, Argatroban, Aspirin, Axitinib, Beniparin, Benzydamine, Bevacizumab, Bismuth, Bivalirudin, Bosutinib, Bromfenac, Cabozantinib, Cangrelor, Caplacizumab, Celecoxib, Cilostazol, Clopidogrel, Cobimetinib, Dabigatran, Dalteparin, Danaparoid, Dasatinib, Dexkeptorofen, Diclofenac, Dipyridamole, Enoxaparin, Epoprostenol, Eptifibatide, Etodolac, Etoricoxib, Flurbiprofen, Heparin, Ibrutanib, Ibuprofen, Iloprost, Imatinib, Indomethacin, Inotersen, Ketoprofen, Ketorolac, Levatinib, Mefenamic acid, Meloxicam, Nabumetone, Naproxen, Nicotinic acid, Nintenanib, Parecoxib, Pazopanib, Phenazone, Phenindione, Piroxicam, Ponatinib, Prasugrel, Regorafenib, Ruxolitinib, Sorafenib, Streptokinase, Sulindac, Sunitinib, Tenecteplase, Tenoxicam, Tiaprofenic acid, Ticagrelor, Tinzaparin, Tirofiban, Tolfenamic acid, Trametinib, Traztuzumab emtansine, Trprostinil, Urokinase, Volanesorsen, Warfarin *(Contraindications to Rivaroxaban)*

6. Renal failure/insufficiency (eGFR<30ml/minute) on the basis of blood investigations (eGFR) within the last 6 months and clinical assessment

7. Severe liver dysfunction on the basis of blood investigations within the last 6 months (liver function and coagulation) and clinical assessment

***Appendix 3:***

***Samples for Future Research and Biobanking Storage***

Approximately 5mL of the research blood taken will be for biobanking at Perspectum and use in future ethically approved research studies research regarding the pathophysiology and mechanism of long COVID, where separate ethical approval may be required. This blood will be stored in an HTA registered biobank at Perspectum’s central laboratory and UCL will remain the custodian.

***Samples for Sub-Study Analysis at Central Laboratories***

Approximately 60 mls of the blood sample will be used for the analyses set out below and relate to the secondary endpoint of the trial. The frozen components of the blood samples will be stored at Perspectum’s central laboratory and sent on to sub-contracted laboratories for testing. In the case of UCLH patient samples for Functional T-cell and live-virus neutralisation antibodies assay, they will be sent direct from the UCLH site to the Francis Crick Institute for analysis.

Guided by clinical practice, patient lived experience and latest scientific hypotheses, the following analyses will be performed in blood samples of some participants, extending to the whole cohort only if there is a clear scientific rationale. Any samples not used within these sub-studies will be retained within the biobank at Perspectum for future research use. All samples will be sent to Perspectum and stored until analysis by third party laboratories:

I.	Genomics analysis: one sample per patient will be taken for initial genome-wide and focused gene (using a long list of immune-regulated genes) analyses. They will be performed using standard protocols adjusting for any population structure. Models will incorporate clinical and environmental determinants of disease severity.

II.	Proteomics analysis one sample per patient. Proteomics will be assessed by proximity extension assay enabling over 1400 proteins to be rapidly analysed. The assay uses oligonucleotide-labelled antibody pairs allowing for pair-wise binding to target proteins.

III.	Metabolomics and Lipidomics; one sample per patient : A combination of Liquid Chromatography with tandem mass spectrometry (LC-MS/MS) based metabolomics and lipidomics will be performed based on a targeted analysis of over 200 metabolites of core metabolism, including acyl-carnitines, acyl-CoAs, amino acids, glycolysis and TCA intermediates and nucleotides using a Thermo Quantiva triple quadrupole mass spectrometer and lipidomics by open-profiling UHPLC-MS/MS using a Thermo Elite Orbitrap interfaced with an Advion Nanomate to allow direct nanoinfusion to detect over 600 annotated lipids.

IV.	Functional T-cell and live-virus neutralisation antibodies in participants recruited to the London trial site. Two samples per patient will be collected (approximately 30mL). Serum and peripheral blood mono-nuclear cells (PBMCs) will be isolated on arrival at the FCI. Cells will be stained and analysed using mass-cytometry and neutralising antibodies quantified in the live-virus neutralisation assay.

V.	Endocrine investigation (thyroid, hypothalamo-pituitary- gonadal and hypothalamo-pituitary- adrenal axes) will be included for those with suggestive symptoms associated with thyroiditis, autoimmune hypothyroidism and adrenal impairment for detailed phenotyping to define potential pathophysiology involved in ongoing organ-specific or physiological abnormalities for example endocrine disturbances explaining diverse nonspecific symptoms, including fatigue, hypothermia and dysmenorrhoea.

***Appendix 4:***

**Functional tests in some Long Covid clinics**

a. Chest X Ray
b. High Resolution Computerised Tomography (CT) scan of the Chest
c. CT Pulmonary Angiogram
d. Pulmonary Function Test
e. 6-minute walk test
f. 1-minute sit to stand test
g. Functional Exhaled Nitric Oxide (FeNO) test
h. Echocardiogram (ECHO)
i. Electrocardiogram (ECG) if the patient had cardiac symptoms
j. Holter monitor of the heart
k. Cardiovascular Magnetic Resonance Scan (CMR)
l. Stress Electrocardiogram
m. Magnetic Resonance Imaging (MRI) scan of Brain
n. Tilt Table Test
o. Coverscan™

***Appendix 5:***

**Functional and Patient Reported Outcome Assessments at Baseline**

All participants must have given consent before the following Patient Reported Outcome Questionnaires and functional tests are carried out:

a. Fatigue Assessment Score(FAS)
b. EuroQol Research Foundation Health related quality of life (EQ-5D-5L)
c. Mental health (GAD-7)
d. Medical Research Council Dyspnoea Score
e. Public Health Questionnaire - Depression (PHQ-9)
f. Perceived Deficit Questionnaire (PDQ-5)
g. Work and Social Adjustment Scale (WSAS) [ Question 4 from Productivity Cost Questionnaire (iPCQ) for absenteeism and Question 8 from iPCQ for presenteeism added]
h. Short Form Questionnaire (SF-12)
i. Cognitive Failure Questionnaire (CFQ) if a patient scores 3 or more on PDQ5 (patients receive an email to complete this questionnaire online via a secure password and patient ID number)
j. Functional abilities and physical function using pedometer monitoring/wearables data
k. Organ impairment and healthcare utilisation
l. Cost-effectiveness of ICP
m. Process outcomes for different ICP components

***Appendix 6:***

**12-Week Assessment Visit**

a. Fatigue Assessment Score
b. IMP accountability (over the phone or in person at the clinic)
c. 6-minute walk test (if performed at baseline visit and where possible undertaken at follow-up)
d. 1-minute Sit to Stand test (if performed at baseline visit and where possible undertaken at follow-up)
e. Medical Research Council (MRC) dyspnoea score
f. Modified Work and Social Adjustment Scale (WSAS) [Q4 from iPCQ for absenteeism and Q8 from iPCQ for presenteeism added]
g. General Anxiety Disorder Questionnaire- 7 (GAD-7)
h. The Primary Care Evaluation of Mental Disorders Patient Health Questionnaire (PHQ-9)
i. EQ-5D
j. Perceived Deficit Questionnaire (PDQ-5)
k. 12-item Short Form Survey (SF12)
l. Cognitive Failure Questionnaire (CFQ) if a patient scores 3 or more on PDQ5 (patients receive an email to complete this questionnaire online via a secure password and patient ID number)
m. Adverse Event review (over the phone or in person in clinic) – Patient completed eCRFs or paper questionnaires will be reported back to site PIs by Lancashire CTU, for review and follow-up of any potential AEs reported by patients
n. Concomitant medication review (over the phone or in person at the clinic)

***Appendix 7:***

**24-Week Assessment Visit**

a. Fatigue Assessment Score
b. IMP accountability (over the phone or in person at the clinic)
c. 6-minute walk test (if performed at baseline visit and where possible undertaken at follow-up)
d. 1-minute Sit to Stand test (if performed at baseline visit and where possible undertaken at follow-up)
e. MRC dyspnoea score
f. Modified Work and Social Adjustment Scale (WSAS) [Q4 from iPCQ for absenteeism and Q8 from iPCQ for presenteeism added]
g. General Anxiety Disorder Questionnaire- 7 (GAD-7)
h. The Primary Care Evaluation of Mental Disorders Patient Health Questionnaire (PHQ-9)
i. EQ-5D-5L
j. Perceived Deficit Questionnaire (PDQ-5)
k. 12-item Short Form Survey (SF12)
l. Cognitive Failure Questionnaire (CFQ), if a patient scores 3 or more on PDQ5 (patients receive an email to complete this questionnaire online via a secure password and patient ID number)
m. Functional ability and Fidelity of delivery of Treatment as Usual and Living with COVID Recovery ^TM^
n. Adverse Event review (over the phone or in person in clinic) for participants on the nested drug trial and participants expressing suicidal ideation on the patient reported outcome questionnaires. Patient completed eCRFs or paper questionnaires will be reported back to site PIs by Lancashire CTU, for review and follow-up of any potential AEs reported by patients. AEs will be reported up to 28 days following the last dose of the trial drugs.
o. Concomitant medication review (over the phone or in person at the clinic) for participants on the nested drug trial only. Concomitant medications will be reported up to 28 days following the last dose of the trial drugs.

***Appendix 8:***

**Data management plan**

Quality Control (QC) includes the operational techniques and activities done within the QA system to verify that the requirements for quality of the trial-related activities are fulfilled. A risk-adapted approach will be used for monitoring. All sites will be centrally monitored for recruitment, data completeness, quality and timeliness of data entry, number of data change requests. A minimum routine remote monitoring schedule of 3 monthly will be set. Any concerns not resolved or raised as a result of the remote monitoring will trigger a full site monitoring visit. To this end:

- Coordinated by LCTU Trial Manager and monitor, site initiation visits will be performed to enable training of local research personnel
- The Trial Manager at LCTU will document the completion of the initiation checklist for each site to verify appropriate approvals are in place prior to initiation of the site
- The Trial Manager will document, as part of the initiation checklist, that all relevant personnel have undergone trial specific training
- Data will be centrally monitored by the LCTU Monitor and Trial Manager to check:
  - Adverse Event reporting rates between centres
  - Screening, recruitment, and dropout rates between centres
  - Data entry consistency. The Data Manager and Trial Manager, with the Monitor, will follow-up on the data queries; Central monitoring reports will thus be generated for the TMG, who will oversee the activity and in accordance with the monitoring plan, will identify when additional intervention, e.g. site visits, should be undertaken.
  - Independent oversight of the trial will be provided by IDMC and independent members of the TSC.

Among the most important factors influencing the delivery of these quality objectives are:

- Minimising the burden on the clinicians working in overstretched LC clinics.
- Ensuring suitability of the participants and having access to the trial treatment without impacting on their other medical needs.
- Ensuring information given to the participants and the PIs in a timely and readily digestible fashion without adversely impacting on the patient care.
- To allow the treating physician to use their clinical judgement to decide whether any of the treatment arms are not suitable for the patient under their care.
- To collect comprehensive information on the mortality as well as morbidity of the LC status.
- In all aspects of the trial, any risks to the patients and well-being will by a key principle in that of proportionality. Risks associated with participation in the trial must be considered in the context of usual care.

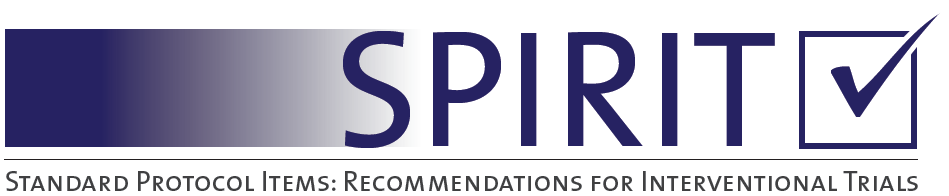

Appendix 8: SPIRIT 2013 Checklist: Recommended items to address in a clinical trial protocol and related documents*

| Section/item | Item No | Description | Addressed on page number |
| --- | --- | --- | --- |
| **Administrative information** | | |  |
| Title | 1 | Descriptive title identifying the study design, population, interventions, and, if applicable, trial acronym | ___1_________ |
| Trial registration | 2a | Trial identifier and registry name. If not yet registered, name of intended registry | ___2_________ |
|  | 2b | All items from the World Health Organization Trial Registration Data Set | __4-14________ |
| Protocol version | 3 | Date and version identifier | ____17_______ |
| Funding | 4 | Sources and types of financial, material, and other support | ___16________ |
| Roles and responsibilities | 5a | Names, affiliations, and roles of protocol contributors | ___1_________ |
|  | 5b | Name and contact information for the trial sponsor | ____16_______ |
|  | 5c | Role of study sponsor and funders, if any, in study design; collection, management, analysis, and interpretation of data; writing of the report; and the decision to submit the report for publication, including whether they will have ultimate authority over any of these activities | ____16_______ |
|  | 5d | Composition, roles, and responsibilities of the coordinating centre, steering committee, endpoint adjudication committee, data management team, and other individuals or groups overseeing the trial, if applicable (see Item 21a for data monitoring committee) | ____16_______ |
| Introduction |  |  |  |
| Background and rationale | 6a | Description of research question and justification for undertaking the trial, including summary of relevant studies (published and unpublished) examining benefits and harms for each intervention | ______3_____ |
|  | 6b | Explanation for choice of comparators | ______3, 11__ |
| Objectives | 7 | Specific objectives or hypotheses | ______3_____ |
| Trial design | 8 | Description of trial design including type of trial (eg, parallel group, crossover, factorial, single group), allocation ratio, and framework (eg, superiority, equivalence, noninferiority, exploratory) | ______4_____ |
| Methods: Participants, interventions, and outcomes | | |  |
| Study setting | 9 | Description of study settings (eg, community clinic, academic hospital) and list of countries where data will be collected. Reference to where list of study sites can be obtained | ______4______ |
| Eligibility criteria | 10 | Inclusion and exclusion criteria for participants. If applicable, eligibility criteria for study centres and individuals who will perform the interventions (eg, surgeons, psychotherapists) | _____6______ |
| Interventions | 11a | Interventions for each group with sufficient detail to allow replication, including how and when they will be administered | _____4-5_____ |
|  | 11b | Criteria for discontinuing or modifying allocated interventions for a given trial participant (eg, drug dose change in response to harms, participant request, or improving/worsening disease) | _____6-7_____ |
|  | 11c | Strategies to improve adherence to intervention protocols, and any procedures for monitoring adherence (eg, drug tablet return, laboratory tests) | ______7_____ |
|  | 11d | Relevant concomitant care and interventions that are permitted or prohibited during the trial | ______6_____ |
| Outcomes | 12 | Primary, secondary, and other outcomes, including the specific measurement variable (eg, systolic blood pressure), analysis metric (eg, change from baseline, final value, time to event), method of aggregation (eg, median, proportion), and time point for each outcome. Explanation of the clinical relevance of chosen efficacy and harm outcomes is strongly recommended | ______8-9_____ |
| Participant timeline | 13 | Time schedule of enrolment, interventions (including any run-ins and washouts), assessments, and visits for participants. A schematic diagram is highly recommended (see Figure) | _____20-21___ |
| Sample size | 14 | Estimated number of participants needed to achieve study objectives and how it was determined, including clinical and statistical assumptions supporting any sample size calculations | __12-13______ |
| Recruitment | 15 | Strategies for achieving adequate participant enrolment to reach target sample size | __5,7,8_______ |
| **Methods: Assignment of interventions (for controlled trials)** | | |  |
| Allocation: |  |  |  |
| 5,6,Sequence generation | 16a | Method of generating the allocation sequence (eg, computer-generated random numbers), and list of any factors for stratification. To reduce predictability of a random sequence, details of any planned restriction (eg, blocking) should be provided in a separate document that is unavailable to those who enrol participants or assign interventions | ___5_________ |
| Allocation concealment mechanism | 16b | Mechanism of implementing the allocation sequence (eg, central telephone; sequentially numbered, opaque, sealed envelopes), describing any steps to conceal the sequence until interventions are assigned | ____5________ |
| Implementation | 16c | Who will generate the allocation sequence, who will enrol participants, and who will assign participants to interventions | _____5_______ |
| Blinding (masking) | 17a | Who will be blinded after assignment to interventions (eg, trial participants, care providers, outcome assessors, data analysts), and how | _____4_______ |
|  | 17b | If blinded, circumstances under which unblinding is permissible, and procedure for revealing a participant’s allocated intervention during the trial | ____N/A______ |
| **Methods: Data collection, management, and analysis** | | |  |
| Data collection methods | 18a | Plans for assessment and collection of outcome, baseline, and other trial data, including any related processes to promote data quality (eg, duplicate measurements, training of assessors) and a description of study instruments (eg, questionnaires, laboratory tests) along with their reliability and validity, if known. Reference to where data collection forms can be found, if not in the protocol | _____6-9______ |
|  | 18b | Plans to promote participant retention and complete follow-up, including list of any outcome data to be collected for participants who discontinue or deviate from intervention protocols | _____10______ |
| Data management | 19 | Plans for data entry, coding, security, and storage, including any related processes to promote data quality (eg, double data entry; range checks for data values). Reference to where details of data management procedures can be found, if not in the protocol | _____32______ |
| Statistical methods | 20a | Statistical methods for analysing primary and secondary outcomes. Reference to where other details of the statistical analysis plan can be found, if not in the protocol | ______10____ |
|  | 20b | Methods for any additional analyses (eg, subgroup and adjusted analyses) | _____11-12___ |
|  | 20c | Definition of analysis population relating to protocol non-adherence (eg, as randomised analysis), and any statistical methods to handle missing data (eg, multiple imputation) | _______11____ |
| **Methods: Monitoring** | | |  |
| Data monitoring | 21a | Composition of data monitoring committee (DMC); summary of its role and reporting structure; statement of whether it is independent from the sponsor and competing interests; and reference to where further details about its charter can be found, if not in the protocol. Alternatively, an explanation of why a DMC is not needed | ____14______ |
|  | 21b | Description of any interim analyses and stopping guidelines, including who will have access to these interim results and make the final decision to terminate the trial | ______12____ |
| Harms | 22 | Plans for collecting, assessing, reporting, and managing solicited and spontaneously reported adverse events and other unintended effects of trial interventions or trial conduct | ____31______ |
| Auditing | 23 | Frequency and procedures for auditing trial conduct, if any, and whether the process will be independent from investigators and the sponsor | ______31____ |
| Ethics and dissemination | | |  |
| Research ethics approval | 24 | Plans for seeking research ethics committee/institutional review board (REC/IRB) approval | ______16_____ |
| Protocol amendments | 25 | Plans for communicating important protocol modifications (eg, changes to eligibility criteria, outcomes, analyses) to relevant parties (eg, investigators, REC/IRBs, trial participants, trial registries, journals, regulators) | __14 ________ |
| Update Consent or assent | 26a | Who will obtain informed consent or assent from potential trial participants or authorised surrogates, and how (see Item 32) | _____5______ |
|  | 26b | Additional consent provisions for collection and use of participant data and biological specimens in ancillary studies, if applicable | ______6-8_____ |
| Confidentiality | 27 | How personal information about potential and enrolled participants will be collected, shared, and maintained in order to protect confidentiality before, during, and after the trial | _______15___ |
| Declaration of interests | 28 | Financial and other competing interests for principal investigators for the overall trial and each study site | ____16_______ |
| Access to data | 29 | Statement of who will have access to the final trial dataset, and disclosure of contractual agreements that limit such access for investigators | _____10_____ |
| Ancillary and post-trial care | 30 | Provisions, if any, for ancillary and post-trial care, and for compensation to those who suffer harm from trial participation | ___N/A______ |
| Dissemination policy | 31a | Plans for investigators and sponsor to communicate trial results to participants, healthcare professionals, the public, and other relevant groups (eg, via publication, reporting in results databases, or other data sharing arrangements), including any publication restrictions | ____2_______ |
|  | 31b | Authorship eligibility guidelines and any intended use of professional writers | ____N/A_____ |
|  | 31c | Plans, if any, for granting public access to the full protocol, participant-level dataset, and statistical code | ____N/A_____ |
| Appendices |  |  |  |
| Informed consent materials | 32 | Model consent form and other related documentation given to participants and authorised surrogates | _Study website__ |
| Biological specimens | 33 | Plans for collection, laboratory evaluation, and storage of biological specimens for genetic or molecular analysis in the current trial and for future use in ancillary studies, if applicable | _____8______ |

*It is strongly recommended that this checklist be read in conjunction with the SPIRIT 2013 Explanation & Elaboration for important clarification on the items. Amendments to the protocol should be tracked and dated. The SPIRIT checklist is copyrighted by the SPIRIT Group under the Creative Commons “[Attribution-NonCommercial-NoDerivs 3.0 Unported](http://www.creativecommons.org/licenses/by-nc-nd/3.0/)” license.
